# The impact of COVID-19 infection experience on risk perception and preventive behaviour: A cohort study

**DOI:** 10.1101/2025.05.07.25327146

**Authors:** Michio Murakami, Mei Yamagata, Asako Miura

## Abstract

**Background:** Understanding how individuals perceive and respond to their own infectious disease experiences offers valuable opportunities to develop effective risk communication strategies. This study examined whether coronavirus disease 2019 (COVID-19) infection experience enhances preventive behaviour, with risk perception acting as a mediating factor.

**Methods:** The study included participants aged ≥18 years residing in Japan, enrolled in a 30-wave cohort study conducted from January 2020 to March 2024. Using propensity score matching, 135 pairs of participants with and without infection were extracted, adjusting for dread and unknown risk perception, preventive behaviours (e.g., hand disinfection and mask-wearing), sociopsychological variables, and individual attributes. Comparisons of risk perception and preventive behaviour were made between groups post-infection experience, and mediation analysis was conducted to test whether risk perception mediated the effect of infection experience on preventive behaviour.

**Results:** No significant differences in covariates were observed between groups prior to infection experience, except gender. Following infection experience, participants in the infection group reported significantly higher scores for one item of unknown risk perception and a greater proportion of mask-wearing, even after adjusting for gender. The indirect effect of infection experience on mask-wearing, mediated by the unknown risk perception item, was significant.

**Conclusion:** COVID-19 infection experience increased perceptions of unknowable exposure, which in turn promoted mask-wearing behaviour. Incorporating insights from personal infection experiences into public health messaging may enhance risk perception and promote preventive behaviour among non-infected individuals, offering a novel approach to infection control at the population level.

## INTRODUCTION

To mitigate the spread of infectious diseases, individual infection preventive behaviours—such as mask-wearing, hand washing, and hand sanitisation—are essential alongside vaccination and broader social regulatory measures, including lockdowns. These individual actions are particularly important due to their relatively low invasiveness and economic cost compared to large-scale interventions. The effectiveness of individual infection prevention has been previously reported.^1, 2^ A systematic review and meta-analysis demonstrated that infection preventive behaviours were effective in slowing the transmission of coronavirus disease 2019 (COVID-19).^3^

Understanding the factors influencing such behaviour is crucial to its promotion. These factors may be classified into contextual factors, socioeconomic position, and intermediary determinants.^4^

Among these, intermediary determinants—such as individual knowledge, attitudes, and risk perception—serve as the direct pathway through which broader structural conditions influence preventive behaviour. In particular, risk perception of infectious diseases is considered a key determinant. Slovic identified two primary dimensions of risk perception: dread and unknown.^5^ Dread risk perception encompasses elements such as high fatality and uncontrollability, whereas unknown risk perception includes characteristics such as a lack of awareness of exposure or the consequences. Stronger perceptions along these dimensions are associated with greater support for regulatory measures. The association between COVID-19 infection preventive behaviours and risk perception has been documented globally.^6^ Furthermore, repeated cross-sectional and cohort studies have observed that COVID-19 risk perception was associated with the implementation of preventive behaviour.^7, 8^ Risk perception of COVID-19 is not constant, with increases after social regulatory measures and decreases after relaxation of measures.^9, 10^

One key determinant of risk perception is prior experience related to the risk itself.^11, 12^ Previous studies have found that individuals who experienced infection personally, within their families, or believed themselves to have been infected, reported higher levels of COVID-19 risk perception.^7, 13^ In addition, cohort studies revealed that individuals with a history of COVID-19 infection were less likely to refrain from social activities such as gatherings with friends or colleagues, whereas those with acquaintances diagnosed with COVID-19 were more likely to restrict such activities.^14^ However, these studies did not control for prior risk perception or preventive behaviour, raising the possibility of reverse causality. As a result, there is limited research examining the causal impact of infection experience on subsequent risk perception and preventive behaviour. A deeper understanding of how infection experience relates to risk perception and preventive behaviour may offer insights into the psychology of individuals who have been infected. In particular, if risk perception mediates the relationship between infection experience and preventive behaviour, recognising differences in risk perception between infected and non-infected individuals could inform public health policy development.

Therefore, this study examined whether experience of COVID-19 infection led to changes in risk perception and preventive behaviours among participants in a panel survey conducted in Japan between January 2020 and March 2024, comprising 30 waves. Using propensity score matching, we selected participants with infection experience and matched them with participants of similar baseline characteristics, including risk perception, preventive behaviours, sociopsychological variables, and individual attributes. We then evaluated the impact of infection experience on risk perception and preventive behaviours, including mask-wearing and hand disinfection. Additionally, we investigated whether infection experience influenced preventive behaviour through the mediating role of risk perception.

## METHODS

### Ethics

This study was approved by the Behavioral Subcommittee of the Research Ethics Committee of the Graduate School of Human Sciences, the University of Osaka (Authorisation Number: HB019-099: until February 2023; HB022-117: from February 2023 onwards). Informed consent was obtained online from all participants prior to the commencement of the survey.

### Participants

The panel survey was conducted among individuals aged ≥18 years residing in Japan. Initially, surveys were administered every two weeks to one month from January 2020, and then every two months from the seventh wave (May 2020), amounting to a total of 30 waves. Detailed descriptions of the survey have been provided in prior studies.^8, 9, 15^ In each wave, inattentive participants were identified using the Directed Questions Scale,^16, 17^ and those who incorrectly answered both questions were excluded. The initial wave included 1,248 participants, and 600 new participants were added during the 13th wave (May 2021) (Figure 1a). Participants received a reward of 120–150 yen per survey.

**Figure 1.**
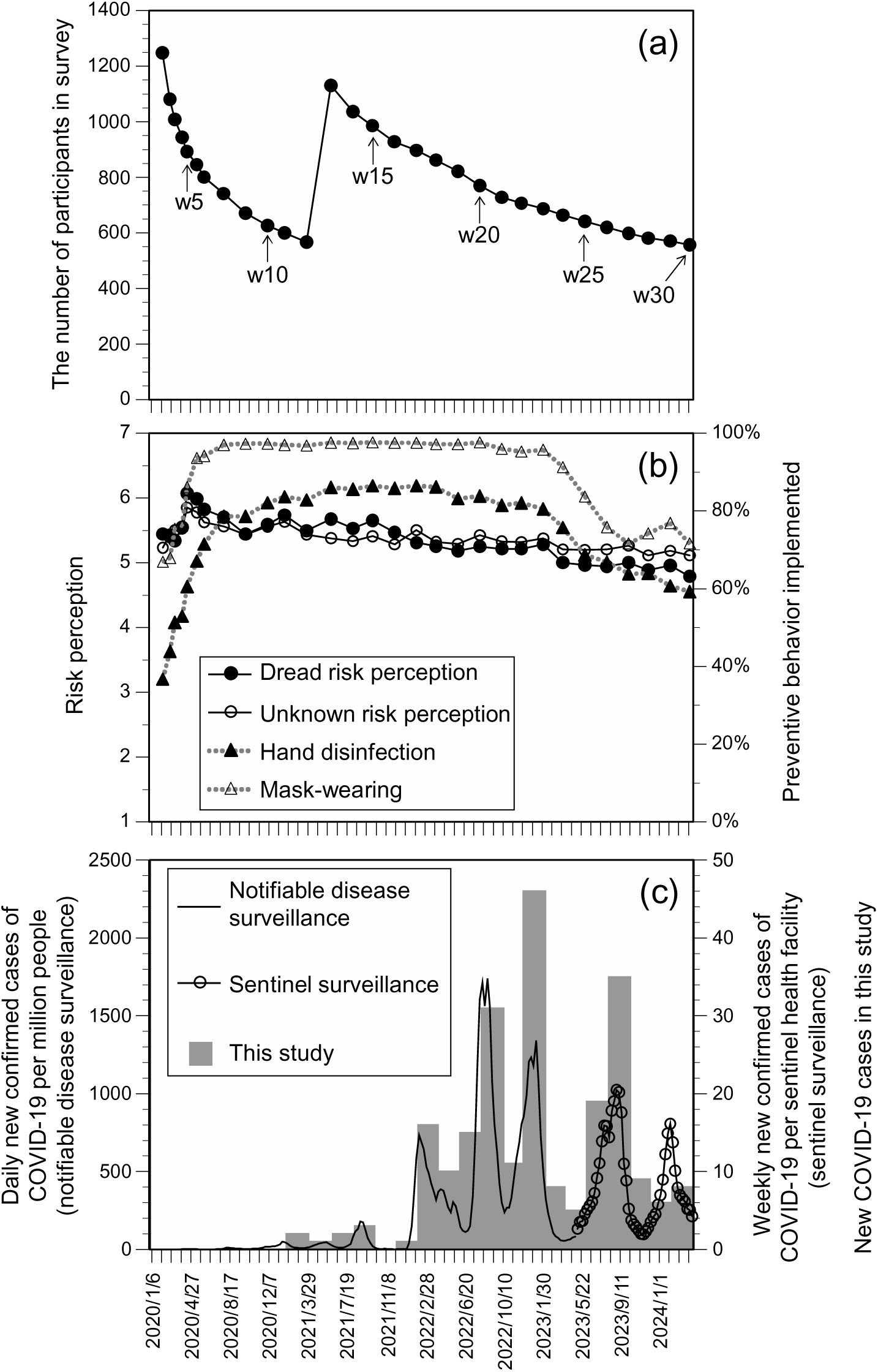
(a) Number of participants per survey wave; (b) trends 389 in risk perception and implementation of preventive behaviour; and (c) number of newly confirmed COVID-19 cases in Japan27, 28 as well as data from this study. In Japan, COVID-19 was reclassified to the same category as seasonal influenza from 8 May 2023, following which preventive measures were relaxed. Notifiable disease surveillance was used before reclassification, and sentinel surveillance thereafter. w: wave.

Questions regarding participants’ own COVID-19 infection experiences were introduced in the 11th wave (January 2021); thus, only participants from wave 11 onwards were included in this study. Of the 1,248 initial participants, 600 remained by the 11th wave, with one individual excluded for not responding to the gender question. Among the 600 new participants in the 13th wave, 11 were excluded for the same reason. Consequently, the total number of participants included in the analysis was 1,188. This exceeded the minimum sample size of 384, as calculated based on the Japanese population, a 95% confidence interval (CI), and a 5% margin of error.^18^

### Survey items

#### Outcome

In line with previous studies,^5, 15^ risk perception related to COVID-19 was assessed using two dimensions: dread and unknown risk. Dread risk perception was evaluated with two items: “Will result in death” (dread risk perception 1) and “Never know when it might happen” (dread risk perception 2). Unknown risk perception was similarly measured using two items: “We may be affected without realising it” (unknown risk perception 1) and “Cannot tell what type of effect this will have” (unknown risk perception 2). Each item was rated on a 7-point Likert scale. The mean scores of the two items were calculated to determine overall dread and unknown risk perceptions, respectively. Additionally, for sensitivity analysis, participants were asked to estimate the probability of COVID-19 infection on a scale from 0 to 100% (hereinafter referred to as the estimated probability of infection).^15^

Infection preventive behaviour was assessed based on whether participants engaged in alcohol-based hand disinfection and mask-wearing.^8^ Temporal changes in risk perception (dread and unknown) and preventive behaviours (hand disinfection and mask-wearing) among the 1,188 participants are depicted in Figure 1b.

#### Exposure factors (infection experience)

A participant’s infection experience was defined based on whether they reported being newly infected in survey wave *x* (i.e. infected between waves *x*−1 and *x*). Participants who indicated infection in wave *x* but not in any prior wave were considered newly infected. As this question was introduced in wave 11, analyses for participants from wave 1 commenced at wave 12, whereas analyses for those joining in wave 13 began at wave 14.

#### Covariates

Covariates included factors associated with infection experience, risk perception, and preventive behaviour in wave *x*, as identified in prior literature.^8, 9, 15, 19–23^ Specifically, items measured in wave *x*-1 were used: dread and unknown risk perception, hand disinfection, mask-wearing, interest in COVID-19 (7-point Likert scale), pathogen-avoidance tendency (mean of five items; 7-point Likert scale),^24, 25^ family members with infection experience, and COVID-19 vaccination history. Gender, age, and residential region (prefectures including Tokyo and designated cities vs. others) were recorded at initial participation (wave 1 or wave 13).

### Statistical analysis

Extraction of individuals in the infection group and non-infection group using propensity score matching

To obtain infection and non-infection groups with similar covariate characteristics but differing in infection experience, participants were selected using propensity score matching.^26^ First, the number of newly infected individuals was confirmed. A total of eight waves were identified—waves *x* = 18–23 (March 2022 to January 2023) and *x* = 26–27 (July to September 2023)—in which ≥10 new infections were reported per wave (Figure 1c). Temporal trends in newly infected individuals generally corresponded with national trends in confirmed COVID-19 cases in Japan.^27, 28^ The infection group comprised participants reporting their first infection in wave *x*, while the non-infection group comprised those without any prior infection experience. Participants with missing values were excluded from propensity score matching due to their small number (Table S1). For each of the eight waves, nearest-neighbour matching was performed with a caliper of 0.01, adjusting for covariates. This resulted in the extraction of 135 matched pairs (270 individuals in total).

Analysis of differences in outcomes and covariates between infected and non-infected individuals

### Analysis 1

To determine whether differences existed in the variance and mean values of covariates between infection and non-infection groups prior to new infection (wave *x*-1), F-tests and t-tests were employed for continuous variables (e.g. risk perception), while chi-square tests were used for categorical variables (e.g. preventive behaviour).

### Analysis 2

To assess whether the outcomes differed between infection and non-infection groups following new infection (wave *x*), F-tests, t-tests, or chi-square tests were conducted, as appropriate.

### Analysis 3

As a significant difference in gender was identified between the two groups in *Analysis 1*, multiple regression and binary logistic regression analyses were performed for risk perception and preventive behaviour, respectively, adjusting for gender.

### Analysis 4

Given that *Analyses 2* and *3* identified significant differences in unknown risk perception 1 and mask-wearing between the two groups, further analysis was conducted using multiple regression and binary logistic regression. In this model, infection experience was treated as the exposure factor, mask-wearing as the outcome, and unknown risk perception 1 as the mediating factor. A 95% CI for the indirect effect was estimated using a bias-corrected method with 2,000 bootstrap samples.

For all *Analyses 1–3*, sensitivity analyses were conducted for each risk perception item and the estimated probability of infection. Analyses were performed using R.^29, 30^ IBM SPSS (version 28), and HAD.^31^ Effect sizes were interpreted following a previous study,^32^ with |d| = 0.20 considered small, 0.50 medium, and |φ| = 0.10 considered small, 0.30 medium. Significant significance was set at *P* = 0.05.

## RESULTS

Comparison of covariates revealed no significant differences, except for gender, between the infection and non-infection groups prior to infection experience (Table 1). A significant difference with a small effect size was noted for gender between the two groups.

**Table 1.** Comparison of covariates between the infection and non-infection groups during the wave preceding infection experience. SD: standard deviation.

| | Average (SD) or n (%) | | P | | | Cohen's<br>d | $\varphi$ |
| --- | --- | --- | --- | --- | --- | --- | --- |
|  | No infection | Infection | F-test | t-test | Chi-test |  |  |
|  | (n = 135) | (n = 135) |  |  |  |  |  |
| Dread risk perception | 5.37 (1.17) | 5.28 (1.11) | 0.523 | 0.521 |  | -0.078 |  |
| Unknown risk perception | 5.41 (1.17) | 5.38 (1.16) | 0.895 | 0.835 |  | -0.025 |  |
| Hand disinfection | 106 (78.5%) | 111 (82.2%) |  |  | 0.444 |  | 0.047 |
| Mask-wearing | 124 (91.9%) | 127 (94.1%) |  |  | 0.475 |  | 0.043 |
| Pathogen-avoidance tendency | 5.42 (1.13) | 5.40 (1.14) | 0.927 | 0.898 |  | -0.016 |  |
| Interest in COVID-19 | 5.59 (1.55) | 5.66 (1.56) | 0.979 | 0.696 |  | 0.048 |  |
| Vaccination | 116 (85.9%) | 115 (85.2%) |  |  | 0.863 |  | -0.011 |
| Infection experience among family | 30 (22.2%) | 31 (23.0%) |  |  | 0.884 |  | 0.009 |
| Age | 39.9 (9.3) | 41.1 (8.3) | 0.068 | 0.290 |  | 0.129 |  |
| Man | 61 (45.2%) | 45 (33.3%) |  |  | 0.046 |  | -0.121 |
| Urban | 105 (77.8%) | 102 (75.6%) |  |  | 0.666 |  | -0.026 |

Regarding outcomes following infection experience (Table 2), no significant differences were observed in the variance or mean values of dread and unknown risk perceptions between the groups. The proportion of participants wearing masks was significantly higher in the infection group than in the non-infection group, whereas no significant difference was observed in the proportion engaging in hand disinfection. The effect size for mask-wearing ranged from small to medium.

**Table 2.** Comparison of risk perception and implementation of preventive behaviour between the infection and non-infection groups in the wave following infection experience. SD: standard deviation.

| | Average (SD) or n (%) | | P | | | Cohen's | $\phi$ |
| --- | --- | --- | --- | --- | --- | --- | --- |
|  | No infection | Infection | F-test | t-test | Chi-test | d |  |
|  | (n = 135) | (n = 135) |  |  |  |  |  |
| Dread risk perception | 5.35 (1.22) | 5.30 (1.18) | 0.565 | 0.723 |  | -0.043 |  |
| Unknown risk perception | 5.39 (1.24) | 5.61 (1.23) | 0.818 | 0.148 |  | 0.177 |  |
| Hand disinfection | 108 (80.0%) | 115 (85.2%) |  |  | 0.261 |  | 0.068 |
| Mask-wearing | 118 (87.4%) | 128 (94.8%) |  |  | 0.032 |  | 0.130 |

Sensitivity analysis showed no significant differences in any items related to risk perception before infection experience between the groups (Table 3). After infection experience, participants in the infection group exhibited significantly higher levels of dread risk perception 2, unknown risk perception 1, and estimated probability of infection compared with the non-infection group. Conversely, dread risk perception 1 was significantly lower in the infection group. Unknown risk perception 2 showed a significant difference in variance but not in mean value. Among the items, unknown risk perception 1 had the highest |d|. Multivariable analyses adjusting for gender yielded similar findings (Table S2).

**Table 3.** Comparison of individual risk perception items between the infection and non-infection groups. SD: standard deviation.

|  |  | Average (SD) |  | P |  | Cohen's d |
| --- | --- | --- | --- | --- | --- | --- |
|  |  | No infection | Infection | F-test | t-test |  |
|  |  | (n = 135) | (n = 135) |  |  |  |
| Before<br>infection<br>experience | Dread risk perception 1: Will result in death | 4.59 (1.73) | 4.32 (1.73) | 0.891 | 0.206 | -0.154 |
|  | Dread risk perception 2: Never know when it might happen | 6.16 (1.06) | 6.24 (0.96) | 0.920 | 0.471 | 0.088 |
|  | Unknown risk perception 1: We may be affected without realising it | 5.87 (1.13) | 5.93 (1.06) | 0.629 | 0.698 | 0.047 |
|  | Unknown risk perception 2: Cannot tell what type of effect this will have | 4.94 (1.79) | 4.83 (1.82) | 0.738 | 0.613 | -0.062 |
|  | Estimated probability of infection | 53.7 (23.8) | 53.7 (24.6) | 0.520 | 0.988 | -0.002 |
| After<br>infection<br>experience | Dread risk perception 1: Will result in death | 4.58 (1.81) | 4.12 (1.88) | 0.704 | 0.041 | -0.249 |
|  | Dread risk perception 2: Never know when it might happen | 6.13 (1.00) | 6.48 (0.99) | 0.791 | 0.004 | 0.358 |
|  | Unknown risk perception 1: We may be affected without realising it | 5.79 (1.24) | 6.33 (0.95) | 0.138 | <0.001 | 0.489 |
|  | Unknown risk perception 2: Cannot tell what type of effect this will have | 6.33 (0.95) | 4.99 (1.76) | 0.031 | 0.651 | -0.055 |
|  | Estimated probability of infection | 54.6 (23.5) | 64.0 (23.5) | 0.488 | 0.001 | 0.398 |

In an analysis with mask-wearing as the outcome, infection experience as the exposure factor, and unknown risk perception 1 as the mediating factor (Table 4), a significant positive association was observed between infection experience and unknown risk perception 1, as well as between unknown risk perception 1 and mask-wearing. The indirect effect of infection experience on mask-wearing, mediated by unknown risk perception 1, was also significant (B [95% CI] = 0.303 [0.126–0.584]).

**Table 4.** Associations between infection experience and mask-wearing, with unknown risk perception 1 (“We may be affected without realising it)” as a mediating factor. Indirect effect of infection experience on mask-wearing via unknown risk perception 1: B (95% CI) = 0.303 (0.126–0.584). B: unstandardised partial regression coefficient; β: standardised partial regression coefficient; CI: confidence interval.

| | $B$ (95%CI) | $\beta$ | $P$ |
| --- | --- | --- | --- |
| Infection experience -> Unknown risk perception 1 | 0.541 (0.278–0.804) | 0.238 | <0.001 |
| Unknown risk perception 1 -> Mask-wearing | 0.561 (0.260–0.863) | 0.323 | <0.001 |
| Infection experience -> Mask-wearing | 0.644 (–0.312–1.600) | 0.163 | 0.186 |

## DISCUSSION

The results of propensity score matching revealed no significant differences between the infection and non-infection groups in terms of risk perception and preventive behaviour prior to infection experience, nor in other sociopsychological variables and individual attributes, except gender. This indicates that the propensity score matching effectively identified comparable pairs of infected and non-infected participants. Following infection experience, the infection group demonstrated higher levels of certain elements of risk perception—specifically, unknown risk perception 1 and estimated probability of infection—as well as increased mask-wearing, compared to the non-infection group. This association remained consistent after adjusting for gender in multivariable analysis. Moreover, an indirect effect of infection experience on mask-wearing, mediated by unknown risk perception 1, was also observed.

Overall, infection experience appears to elevate unknown risk perception—that is, the belief that one might be infected without realising it—which in turn promotes mask-wearing. Although estimated probability of infection is a distinct construct from dread or unknown risk perception, its increase post-infection is consistent with heightened perception of potential undetected infection.

These findings suggest that infection experience heightens the perceived risk of unrecognised infection, resulting in a higher estimated probability of infection. In reality, however, infection provides a degree of immunity, so the actual risk of reinfection does not increase among individuals who have experienced COVID-19 infection.^33^ In this study, infected participants were identified from the 18th survey wave (March 2022), which corresponded with the spread of the Omicron variant.^34^ The high transmissibility of Omicron may have contributed to a perception among infected participants that they had been unknowingly infected.

Conversely, this study found that infection experience had an ambivalent effect on dread risk perception items 1 and 2. While dread risk perception 2—the fear of not knowing when infection might occur—increased similarly to unknown risk perception 1, dread risk perception 1—the fear that COVID-19 could be fatal—decreased. A previous study reported that individuals who had experienced severe COVID-19 themselves, or in their families, exhibited heightened fear of the disease between 2020 and 2022.^13^ One explanation for the differing results may be the timing of this study, which was conducted after the spread of the Omicron variant, known for reduced severity.^35^ In addition, few older adults (aged ≥65 years)^36^—those at higher risk of severe illness—were included in the study sample (mean ± standard deviation: 41.1 ± 8.3 years old for the infection group and 39.9 ± 9.3 years old for the non-infection group [Table 1]). The participants also consisted of individuals who had survived the infection and were able to complete the surveys. Although no significant association was found between infection experience and hand disinfection, the increase in its practice following infection mirrored the trend seen in mask-wearing.

Importantly, the rise in unknown risk perception and estimated probability of infection may encourage greater self-protective behaviour. A previous study conducted in Japan in February 2021 reported that individuals with prior infection did not refrain from social gatherings, whereas those with infected friends did.^14^ However, that study did not account for pre-infection social activity levels, suggesting that the direction of causality may be reversed—individuals who did not reduce social interaction may have been more likely to contract the virus. Another key difference between the studies is the timing of data collection. As previously noted, this study was conducted following the emergence of the more widespread Omicron variant. Furthermore, after the vaccination campaign began in February 2021, vaccinated individuals increased their preventive behaviour until September 2021.^37^ Taken together, these studies suggest that individuals do not necessarily reduce preventive behaviour after acquiring immunity; on the contrary, they may reinforce it. This study offers valuable insights into how shifts in infection risk perception following infection experience can influence preventive behaviour.

The findings indicate that individuals who have experienced infection develop a distinct risk perception compared to those who have not. Therefore, sharing the personal insights and risk perceptions of infected individuals with those who remain uninfected could foster greater preventive behaviour among the wider public. Disseminating such personal narratives through public information media—while taking care to avoid stigmatisation—may serve as an effective risk communication strategy to promote a prevention-focused mindset in society.

This study has several limitations. First, as it utilised a cohort of online survey participants, selection bias may have occurred in the recruitment of respondents and through survey attrition. Caution should therefore be exercised in generalising the findings. Second, infection status was based on self-reported responses. However, as infection was assessed in each survey wave, recall bias was minimized. Additionally, the number of newly infected participants in this study aligned with nationwide infection trends in Japan. Third, data on infection routes and severity were not collected. Fourth, although the infection and non-infection groups were formed using propensity score matching, the absence of random assignment limits the ability to infer causality.

## CONCLUSION

Infection experience increased unknown risk perception, particularly the belief that one may be affected without realising it. It also led to greater adoption of mask-wearing, with unknown risk perception mediating this relationship. Sharing the experiential insights and risk perceptions of infected individuals with those who have not been infected may be a promising strategy for enhancing infection prevention efforts in public contexts.

## Data Availability

All data produced in the present study are available upon reasonable request to the authors.

## Acknowledgements

We would like to thank Editage (www.editage.com) for English language editing. This work was supported by the Human Science Project, Graduate School of Human Sciences, The University of Osaka, and “The Nippon Foundation- Osaka University Project for Infectious Disease Prevention”, and JSPS KAKENHI Grant Number JP24K21500.

