## Supplementary material for "The impact of COVID-19 infection experience on risk perception and preventive behaviour: A cohort study": Tables S1 and S2

Table S1. Number of participants excluded, included, and retained in the propensity score matching analysis.

| wave *x* | Participants in wave *x* | Participants with infection experience before wave *x*-1 | Participants without infection experience before wave *x*-1 and with missing data in outcome, exposure or covariates ^a^ | Participants for propensity score matching | Extracted pairs (participants) |
| --- | --- | --- | --- | --- | --- |
| 18 | 853 | 10 | 0 | 843 | 11 (22) |
| 19 | 813 | 25 | 0 | 788 | 9 (18) |
| 20 | 761 | 32 | 0 | 729 | 10 (20) |
| 21 | 719 | 45 | 0 | 674 | 27 (54) |
| 22 | 698 | 74 | 14 | 610 | 7 (14) |
| 23 | 678 | 81 | 14 | 583 | 30 (60) |
| 26 | 613 | 119 | 14 | 480 | 13 (26) |
| 27 | 592 | 135 | 11 | 446 | 28 (56) |

Table S2. Associations between infection experience and both risk perception and preventive behaviour, adjusted for gender. B: unstandardised partial regression coefficient; OR: odds ratio; CI: confidence interval. Dread risk perception 1: Will result in death; Dread risk perception 2: Never know when it might happen; Unknown risk perception 1: We may be affected without realising it; Unknown risk perception 2: Cannot tell what type of effect this will have.

|  | B (95%CI) | | | | | | | OR (95%CI) | |
| --- | --- | --- | --- | --- | --- | --- | --- | --- | --- |
|  | Dread risk perception | Unknown perception | Dread risk perception 1 | Dread risk perception 2 | Unknown risk perception 1 | Unknown risk perception 2 | Estimated probability of infection | Hand disinfection | Mask-wearing |
| Infection experience | −0.064  (−0.355–0.227) | 0.199  (−0.099–0.497) | −0.466  (−0.911–−0.021) | 0.338  (0.098–0.578) | 0.531  (0.263–0.798) | −0.133  (−0.587–0.321) | 8.94  (3.27–14.60) | 1.33  (0.70–2.54) | 2.64  (1.05–6.62) |
| Man | −0.102  (−0.399–0.196) | −0.165  (−0.470–0.141) | −0.056  (−0.512–0.400) | −0.147  (−0.393–0.098) | −0.084  (−0.358–0.190) | −0.245  (−0.710–0.220) | −3.54  (−9.34–2.26) | 0.52  (0.27–0.98) | 1.00  (0.42–2.38) |
